# Individual and Occupational Predictors of Sleep Intervention Response Among Shift-Working Firefighters

**DOI:** 10.1101/2025.10.27.25338848

**Authors:** Yun Min Song, Saebom Jeon, Ayeong Cho, Seockhoon Chung, Yeonsoon Ahn, Sooyeon Suh, Jae Kyoung Kim

## Abstract

**Background:** Cognitive Behavioral Therapy for Insomnia (CBT-I) is the first-line treatment for chronic insomnia, yet its effectiveness varies substantially across individuals. This variability is particularly pronounced in shift workers, who often experience irregular sleep-wake cycles and high levels of stress.

**Objective:** We examined heterogeneous CBT-I response patterns and their predictors in a high-risk shift-working firefighter cohort, with the goal of identifying modifiable clinical and occupational factors that can inform personalized intervention strategies.

**Methods:** Over a five-week CBT-I intervention, self-reported sleep quality was longitudinally assessed in 50 participants (mean age = 34.8 ± 7.7 years; 88% male). Growth mixture modeling identified latent response trajectories, and multinomial logistic regression examined predictors including sleep parameters, insomnia severity, alcohol use, and work-related characteristics. A validated mathematical alertness model was used to objectively characterize physiological response patterns.

**Results:** Three response patterns emerged: steady-poor (24%), steady-moderate (50%), and improving (26%). Poor response was associated with emergency duty, longer sleep onset latency, and higher insomnia severity. Improved response was predicted by 3-day shift cycles, lower alcohol intake, and shorter shift-work duration. Mathematical model–predicted alertness profiles paralleled subjective response trajectories.

**Conclusions:** CBT-I response among shift workers is highly heterogeneous and shaped by both clinical and occupational factors. Identifying these moderators can guide tailored behavioral interventions to improve treatment outcomes for individuals with insomnia in demanding work environments.

## 1. Introduction

Cognitive-behavioral therapy for insomnia (CBTI) is considered the first line of treatment by many reputable organizations (Qaseem et al., 2016; Riemann et al., 2023; Siebern & Manber, 2011). However, its effects have been reported to be limited for shift workers; a recent meta-analysis found no clinically significant improvements post-CBTI for shift workers (Reynolds et al., 2023). The complexity of investigating this diverse population introduces challenges in tailoring interventions, as it involves work-related factors such as shift cycle, characteristics of shift work (emergency response vs. fire suppression), quick returns, and individual factors such as sleep-related behaviors and lifestyle factors. While several studies have investigated week-to-week changes during CBTI to assess factors, such as the rate of change or to identify the timing of frequent drop-outs (Peter et al., 2019; Sweetman et al., 2020), none have yet delved into these changes to uncover predictors of divergent treatment outcomes among shift workers.

As a treatment outcome, sleep quality is important for intervention as it is associated with various factors, including physical and mental health (Jaradat et al., 2020; Lo et al., 2018). A meta-analysis reported a dose-response relationship between sleep quality and mental health, indicating that greater improvements in sleep were associated with greater improvements in mental health (Scott et al., 2021). For shift workers, poor sleep quality is associated with a range of risks, including physiological and psychological disorders such as metabolic syndrome and depression (Demiralp & Ozel, 2021; Lee et al., 2023). Many studies evaluate sleep quality as an important outcome variable in sleep interventions for shift workers (Crowther et al., 2021). However, assessing sleep quality in shift workers is challenging due to the day-to-day variability of sleep caused by shift work.

Previous studies have primarily utilized cross-sectional or pre-post study designs (Alves et al., 2023; Jeong et al., 2019; Savall et al., 2021), which hampered the exploration of long-term changes in intervention effects. Few studies have investigated individual variations in sleep patterns among shift workers over time as they participate in CBTI, while work-related characteristics likely influence the sleep quality of shift workers. Furthermore, although predictors of treatment outcomes have been extensively studied (Huang & Huang, 2023; Monma et al., 2018), their associations with different sleep quality trajectories during the intervention remain unclear. Thus, adopting a multi-wave design, which offers a dynamic perspective at each stage of the intervention period, may provide clinical insights for future studies. For instance, it would allow us to discern when changes occur during the intervention and whether the trajectory of improvement occurs in the initial stages of intervention or after an extended duration. Additionally, it would facilitate the identification of characteristics associated with distinct trajectories, thereby identifying characteristics of individuals who may benefit most, as well as other factors that warrant additional attention.

To the best of our knowledge, no study has explored evolving patterns of sleep quality using a person-centered approach among shift workers participating in CBTI. In addition, the relationships between factors such as working conditions (e.g., shift cycles, primary duty, and shift work period) and individual characteristics (e.g., demographics, alcohol consumption, insomnia symptoms) with different sleep quality trajectories remain unclear.

Previous studies have shown that sleep quality is associated with personal characteristics. Studies in the general adult population have found demographic and lifestyle factors, such as gender, BMI, and nutritional status, related to sleep quality (İçer & Gezmen Karadağ, 2023; Madrid-Valero et al., 2017). For shift-working nurses, irregular shift cycles have been shown to impact sleep quality negatively (Albakri et al., 2024). Based on the existing theories and research findings, we hypothesize that: (1) there will be at least two or more latent patterns in sleep quality trajectories among shift workers undergoing intervention; (2) the latent patterns will be associated with work-related factors, such as irregular shift cycles, more frequent emergency calls, and longer shift durations; and (3) the latent patterns will also be associated with individual characteristics, such as demographics, lifestyle factors, and insomnia severity.

To investigate this, we analyze daily subjective sleep quality (How is your sleep quality today?) rated on a scale of 1-5 (SQ) collected from shift-working firefighters undergoing a five-week CBTI intervention. Previous studies using the same assessment have explored the relationship between subjective daily sleep quality and other factors, revealing positive associations with positive affect and exercise behavior (Dzierzewski et al., 2014; McCrae et al., 2008). Additionally, improved sleep quality was associated with total sleep time, and impaired sleep quality was associated with time in bed (Garefelt et al., 2022). Through this analysis, we aim to explore the longitudinal changes in individual SQ throughout the intervention, identify diverse latent patterns in the trajectories, and examine the key factors that differentiate these patterns (Fig 1).

**Figure 1.**
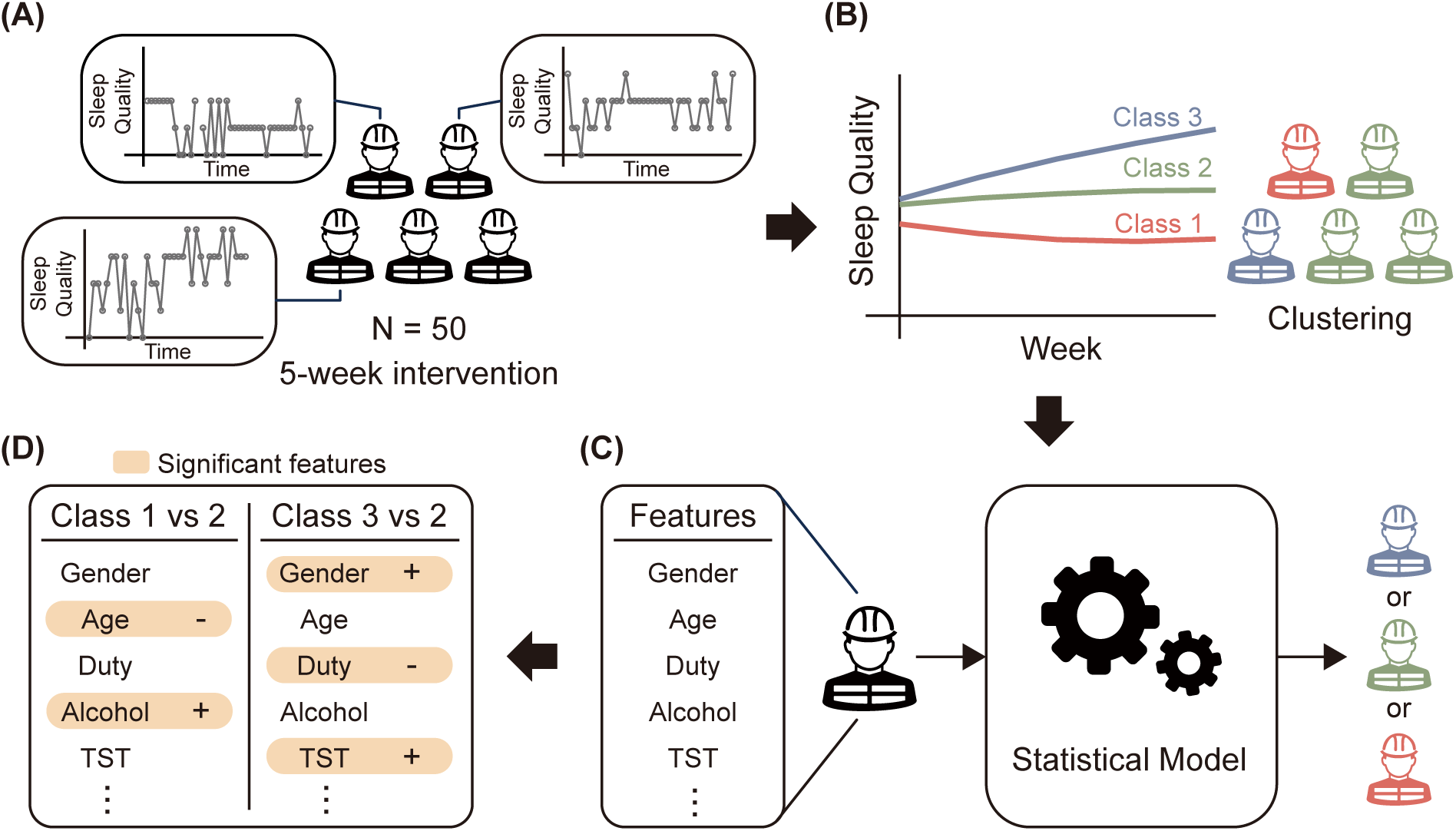
Study overview. (A) Daily time series data on self-reported sleep quality were collected for each of the 50 firefighters participating in weekly cognitive behavioral therapy for insomnia. The daily time series of the 50 participants exhibited diverse trends, variations, observation dates, and numbers of observations, posing challenges in assessing the intervention’s effectiveness. (B) To address this issue, we identified distinct classes within the collected time series that share similar weekly variations throughout the intervention period using a growth mixture model. (C) Subsequently, we employed a statistical model to predict the latent trajectory classes based on participants’ demographic, lifestyle, and clinical information and sleep features. (D) The statistical model identified features that significantly influence the effectiveness of the intervention.

## 2. Methods

### 2.1. Participants

Participants in this study were recruited to participate in FIT-INdividual (Firefighter’s Therapy for Insomnia and Nightmares-individual version), a sleep intervention tailored for firefighters in South Korea (see **Appendix A** for details). This intervention was available exclusively to shift-working firefighters experiencing sleep problems. Participants were recruited through advertisements at each fire station in Korea. They received a five-week sleep intervention program and a sleep report containing detailed information about their sleep patterns.

The intervention was conducted in 2022 for shift-working firefighters at 26 locations in a metropolitan district (Gyeonggi-do) that was in the process of transitioning from the 21-day cycle to the 3-day cycle at the time of the intervention, to explore the effect of the shift-working cycle. All firefighters working shifts at that time were eligible to participate as long as they were experiencing sleep problems, so there were no exclusion criteria for specific sleep disorders or psychiatric disorders. A total of 60 participants were initially recruited, though four individuals withdrew from the program due to personal reasons or perceived burdens associated with attending weekly sessions. Due to insufficient data collection resulting from non-compliance with completing writing sleep diaries and wearing actigraphy, four additional participants were excluded. Two additional participants were excluded during the pre-processing stage of the analyses, resulting in a sample of 50 participants.

All methods and procedures of this study were in accordance with the Declaration of Helsinki and were approved by the Institutional Review Board of Yonsei University Wonju Severance Christian Hospital (CR318031). Participants were informed of the nature and purpose of the study, and all signed informed consent forms.

### 2.2. Data collection

Prior to the intervention, participants volunteered for the study using an online form that collected demographic, lifestyle, and work-related information. They also completed self-reported questionnaires on mental health and sleep before and after the intervention. The online form was constructed and collected through an online platform (Google Forms). Participants were instructed to wear an actigraphy device continuously and maintain a weekly sleep diary throughout the five-week program. These tools were used to track changes in various aspects of sleep, including sleep patterns and SQ. Data collection spanned a total of five weeks, from the week before the first session to the week following the completion of the fourth session. Detailed descriptions of the measures and data preprocessing procedures are provided in **Appendix B**.

### 2.3. Analysis

To assess individual differences in SQ trajectories during the five-week intervention period, we conducted a time series clustering analysis using growth mixture modeling (GMM; see **Appendix C.1** for details). Specifically, SQ data were used to compute average SQ for each week, allowing for identification of latent trajectory classes across five time points. To explore predictors of class membership, we performed multinomial logistic regression using demographic, lifestyle, clinical, and sleep and work-related variables, with backward elimination employed to address multicollinearity (see **Appendix C.2** for details). Additionally, a validated mathematical model of alertness was applied to simulate individual alertness prediction profiles based on sleep patterns derived from actigraphy and sleep diaries (see **Appendix C.3** for details). These model-derived profiles serve as an indirect objective measure of the intervention’s effects.

## 3. Results

### 3.1. The growth mixture model approach revealed the existence of the three latent classes within the time series

A total of 50 participants completed the intervention programme on their own schedule, with a mean duration of 37.12±7.20 days (see Table 1 for participant demographic and lifestyle information). The whole periods were separated into five periods of baseline days before the first session (week 1, 8.24±1.57 days), between sessions (weeks 2, 7.48±2.21 days; week 3, 7.24±2.67 days; and week 4, 7.40±2.62 days), and the period after the last session (week 5; 6.76± 1.73). The collected SQ time series for each of the participants showed different trends and variations, as well as different observation dates and inter-intervention periods, which posed a challenge in determining the intervention effect. To assess the intervention effect over time, we averaged the measured SQ values for each week, resulting in weekly time series with five data points (Fig 2A bottom).

**Figure 2.**
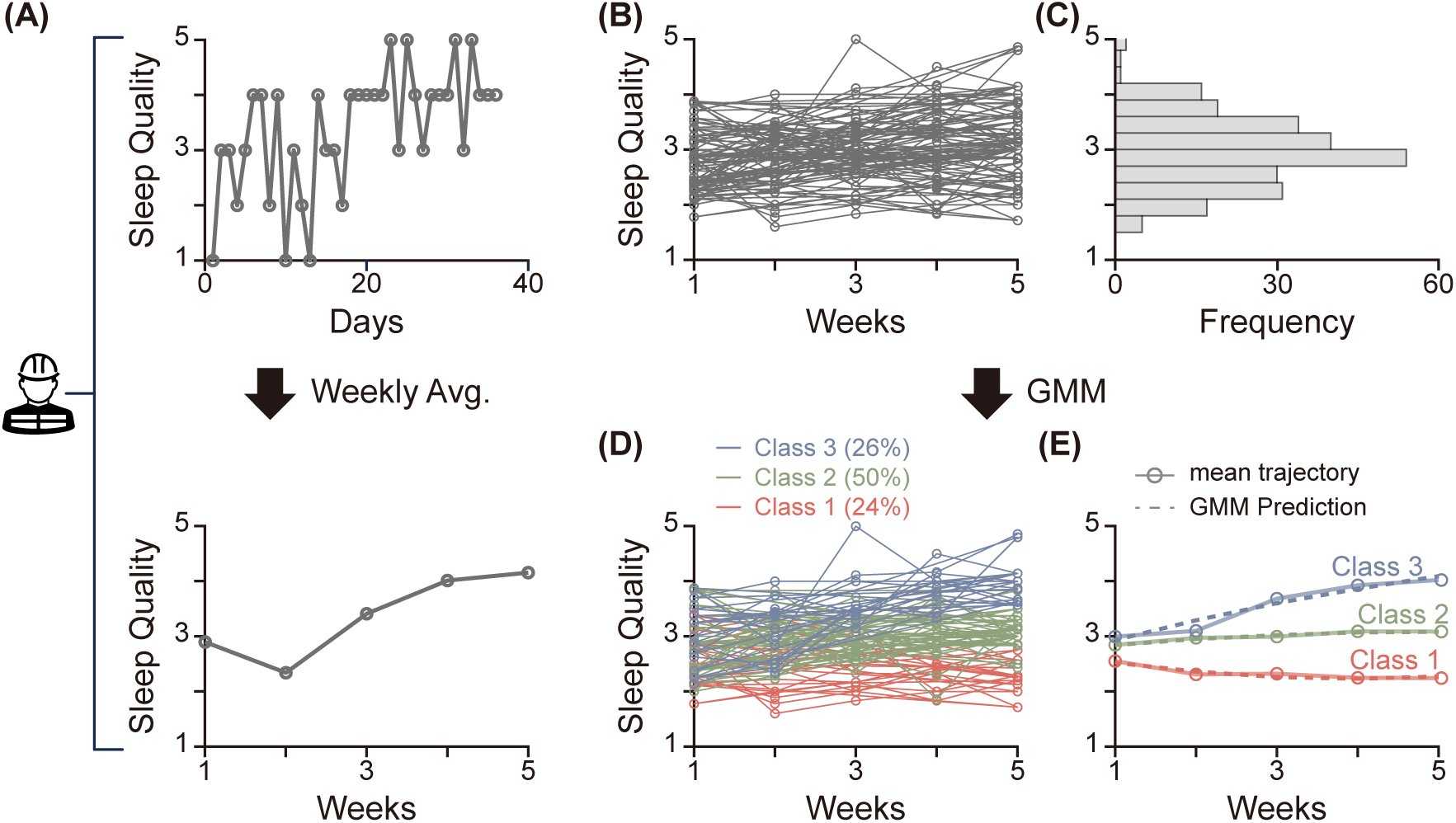
The growth mixture model (GMM) approach revealed three latent classes within the sleep quality time series. (A) Each participant’s daily self-reported sleep quality (rated 1 to 5) time series spanning approximately five weeks (top) is aggregated weekly, resulting in a time series with five-time points (bottom). (B) The weekly time series for 50 participants appears clumped together, displaying a subtle improving pattern over time. (C) Weekly averaged sleep quality follows a normal distribution with a mean of 2.98 and a standard deviation of 0.64. (D) The optimal GMM with the maximum entropy value (see **Methods** for details) classified the 50 time series into three classes (Class 1-3), comprising 24%, 50%, and 26% of the total membership, respectively. (E) The GMM predictions (dashed lines) accurately capture the mean trajectory (solid lines with circles) of the time series in each class, showing patterns of steady low levels (Class 1), steady moderate levels (Class 2), and improving levels (Class 3).

**Table 1.**
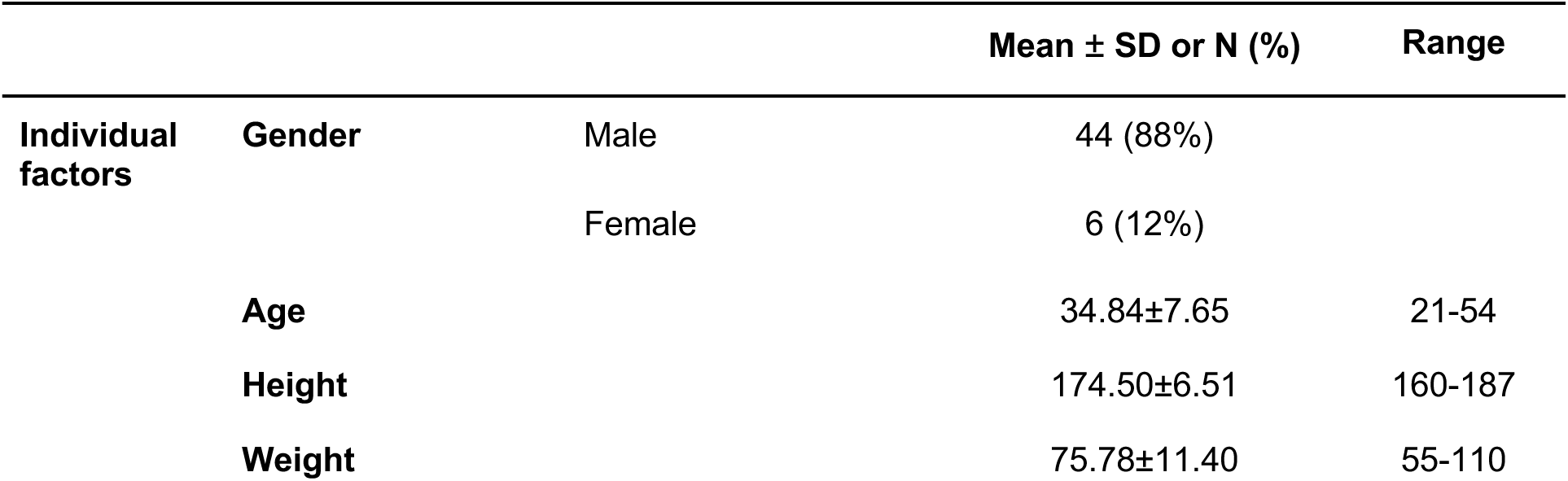
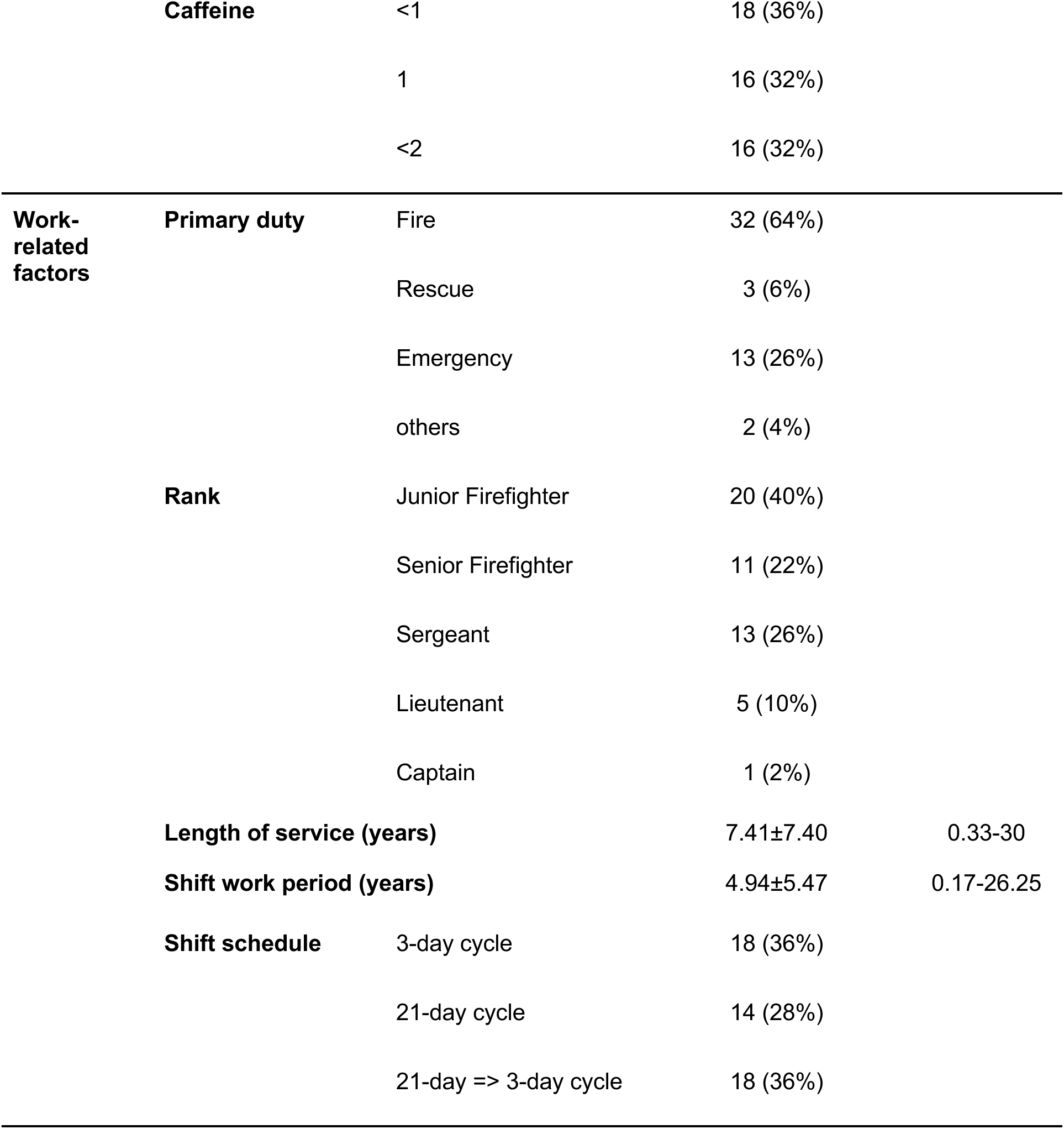
Demographic and lifestyle information of the 50 participants.

The aggregated weekly time series of all participants displayed a slightly increasing trend, suggesting improved SQ associated with the intervention (Fig 2B). However, individual time series exhibited diverse patterns, with a few individuals even showing decreasing trends over time. In order to identify latent classes with similar trends, GMM was applied to the time series data. The GMM, which requires normality, was applicable since SQ exhibited a normal distribution (Fig 2C). (see Methods for details). The results indicated three latent classes underlying the time series group (Class 1–3; Fig 2D), which comprised 24%, 50%, and 26% of the total participants, respectively. Participants in each class exhibited patterns of consistently low SQ (Class 1), moderate SQ (Class 2), and improving SQ (Class 3), respectively, as captured by the mean trajectory of each class (Fig 2e, solid lines). The mean trajectories were closely aligned with the GMM predictions for each class, with statistically significant intercepts estimated as 2.75, 2.69, and 2.54, respectively (Fig 2E, dashed lines). Only the GMM prediction of Class 3 has a statistically significant non-zero slope of 0.42. Taken together, participants could be categorized into three groups displaying distinct responses to the intervention: those with consistently poor SQ, those with consistently moderate SQ, and those experiencing improved SQ.

### 3.2. The multinomial logistic model approach identified six key features influencing sleep quality patterns

We identified three distinct patterns of treatment response in SQ. This prompted an investigation into the underlying factors driving these variations. We built a comprehensive prediction model of latent class subgroups (n=243) using 14 demographic, lifestyle, and clinical variables and 6 sleep parameters that may influence SQ (Fig 3A). Demographic and lifestyle variables include individual factors, including gender, age, caffeine/alcohol intake, in addition to work-related factors such as primary duty, rank, shift schedule, shift work period, and number of dispatches. Clinical variables include Insomnia Severity Index (ISI), Fatigue Severity Scale (FSS) and Shift Work Disorder Questionnaire (SWDQ) scores observed before the intervention to account for individual baseline levels. Sleep parameters include weekly averaged total sleep time (TST), sleep efficiency (SE), number of awakenings (NWAK), wakefulness after sleep onset (WASO), sleep onset latency (SOL), and number of naps (Nap). See Methods for further details of the variables.

**Figure 3.**
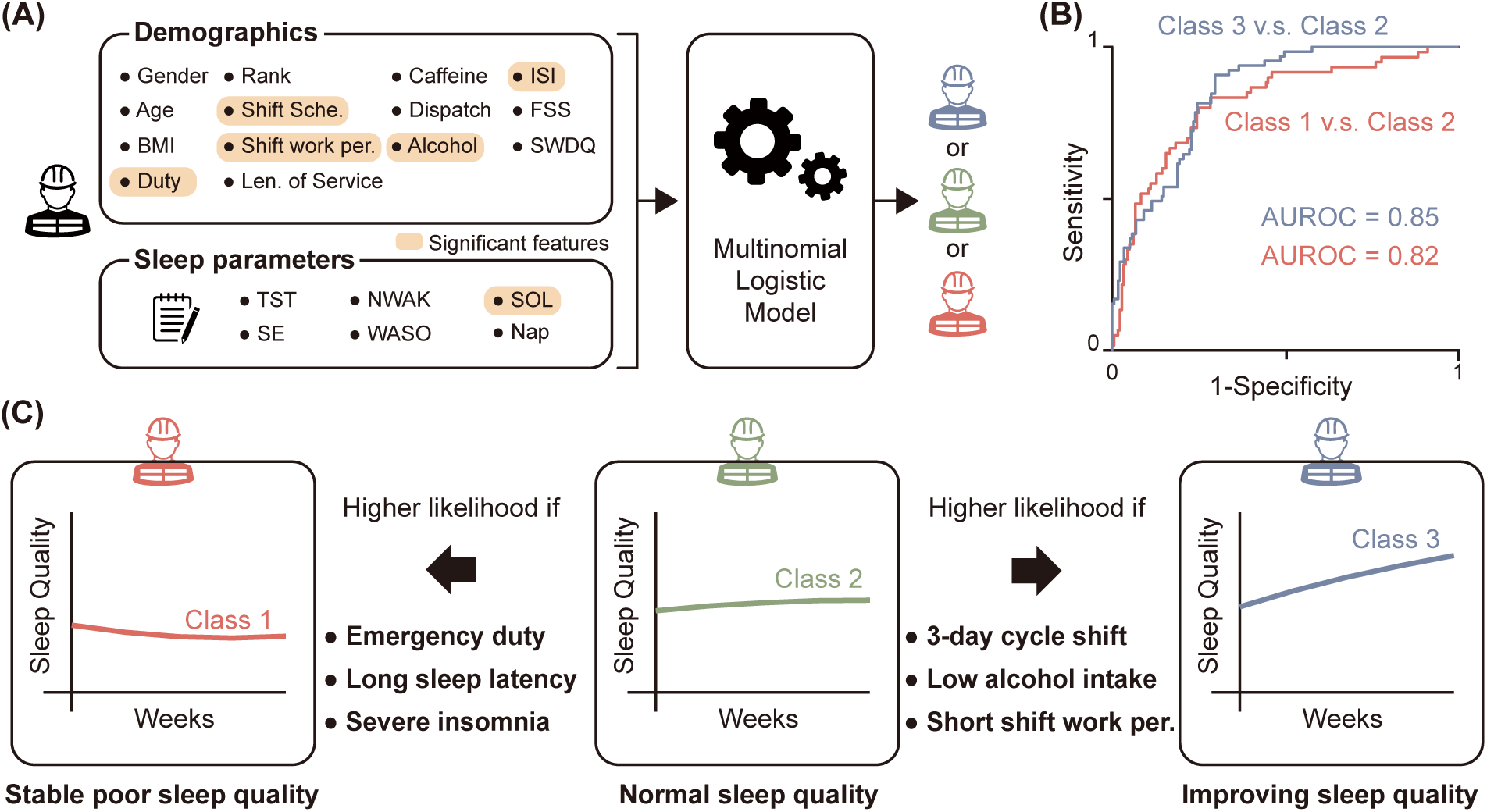
The multinomial logistic model (MLM) approach identifies six key features influencing the sleep quality patterns during the intervention. (A) A multinomial logistic model was fitted to classify each participant into the three identified classes based on 14 demographic, lifestyle, and clinical features and 6 daily sleep parameters collected via sleep diaries. Out of the 20 variables, only 6 were found to be significant in the MLM model. (B) The MLM model accurately predicts the participants’ class with AUROC of 0.85 and 0.82 when classifying Class 3 and 2 and Class 1 and 2, respectively. (C) Analysis of the MLM model revealed that emergency duty, long sleep onset latency, and severe insomnia increase the likelihood of being in Class 1, characterized by steady poor sleep quality, compared to Class 2, which represents steady moderate sleep quality. Conversely, a 3-day rather than 21-day cycle shift, low alcohol intake, and short shift work periods increase the likelihood of being in Class 3, indicative of improving sleep quality.

Utilizing these features, we employed a multinomial logistic model (MLM) to predict each participant’s class (see Methods for details). While fitting the model, backward elimination was used for variable selection to avoid multicollinearity. Consequently, the model retained seven predictors (see Table 2 for details), achieving an AUROC of 0.82 for distinguishing Class 1 from Class 2 and 0.85 for distinguishing Class 3 from Class 2 (Fig 3B). Of the seven retained predictors, six features were statistically significant at the 0.05 level (p-value < 0.05) (Fig 3A, labeled variables). Specifically, Duty, SOL, and ISI emerged as significant factors distinguishing Class 1 (poor SQ group) from 2, while Shift Work Schedule, Shift Work Period, and Alcohol consumption were significant in distinguishing Class 3 (SQ improving group) from 2. The odds ratios calculated for these predictors yielded insightful conclusions (Table 2 and Fig 3C): Emergency duty, a one-minute increase in SOL, and a one-point increase in ISI were associated with a 3.19-, 1.05-, and 3.03-fold greater likelihood of persistent poor SQ, respectively, rather than steady normal sleep. Conversely, adherence to a 3-day cycle shift work schedule was linked to a 4.97-fold higher likelihood of improved SQ, while each additional weekly alcohol intake and year of shift work period decreased this likelihood by 0.40- and 0.76-fold, respectively.

**Table 2.** Predictors of the fitted multinomial logistic model and their odd ratios.

| Variables | Class 1 vs Class 2 |  |  | Class 3 vs Class 2 |  |  |
| --- | --- | --- | --- | --- | --- | --- |
|  | OR | Lower CI | Upper CI | OR | Lower CI | Upper CI |
| Gender | 0.334* | 0.107 | 1.048 | 0.483 | 0.133 | 1.758 |
| Alcohol | 0.894 | 0.666 | 1.199 | 0.404*** | 0.276 | 0.59 |
| SOL | 1.054*** | 1.019 | 1.09 | 0.995 | 0.957 | 1.034 |
| ISI | 3.034*** | 1.503 | 6.125 | 1.029 | 0.518 | 2.041 |
| Duty | 3.190*** | 1.337 | 7.611 | 2.12 | 0.699 | 6.431 |
| Shift Schedule | 2.131 | 0.938 | 4.84 | 4.968*** | 1.913 | 12.899 |
| Shift work period | 1.058 | 0.976 | 1.147 | 0.758*** | 0.656 | 0.876 |

\* $p < 0.1$ , \*\* $p < 0.05$ , \*\*\* $p < 0.01$

### 3.3. The identified classes’ model-predicted alertness shows similar patterns with sleep quality

A formal analysis relies solely on subjective sleep quality evaluation (i.e., SQ), raising the question of whether an objective assessment of the intervention is feasible. To address this, we applied a mathematical model that predicts temporal changes in alertness (AL) based on individual sleep patterns (see Fig 4A and **Appendix C.3** for details). We calculated the mean AL level during the entire wake period (white region in Fig 4A) for each week (AL_w_), excluding the first week due to potential prediction errors resulting from inaccurate initial conditions. A higher AL_w_ represents less sleepiness experienced during wake periods (Song et al., 2023) and proper alignment of sleep periods with the circadian rhythm (Hong et al., 2021).

**Figure 4.**
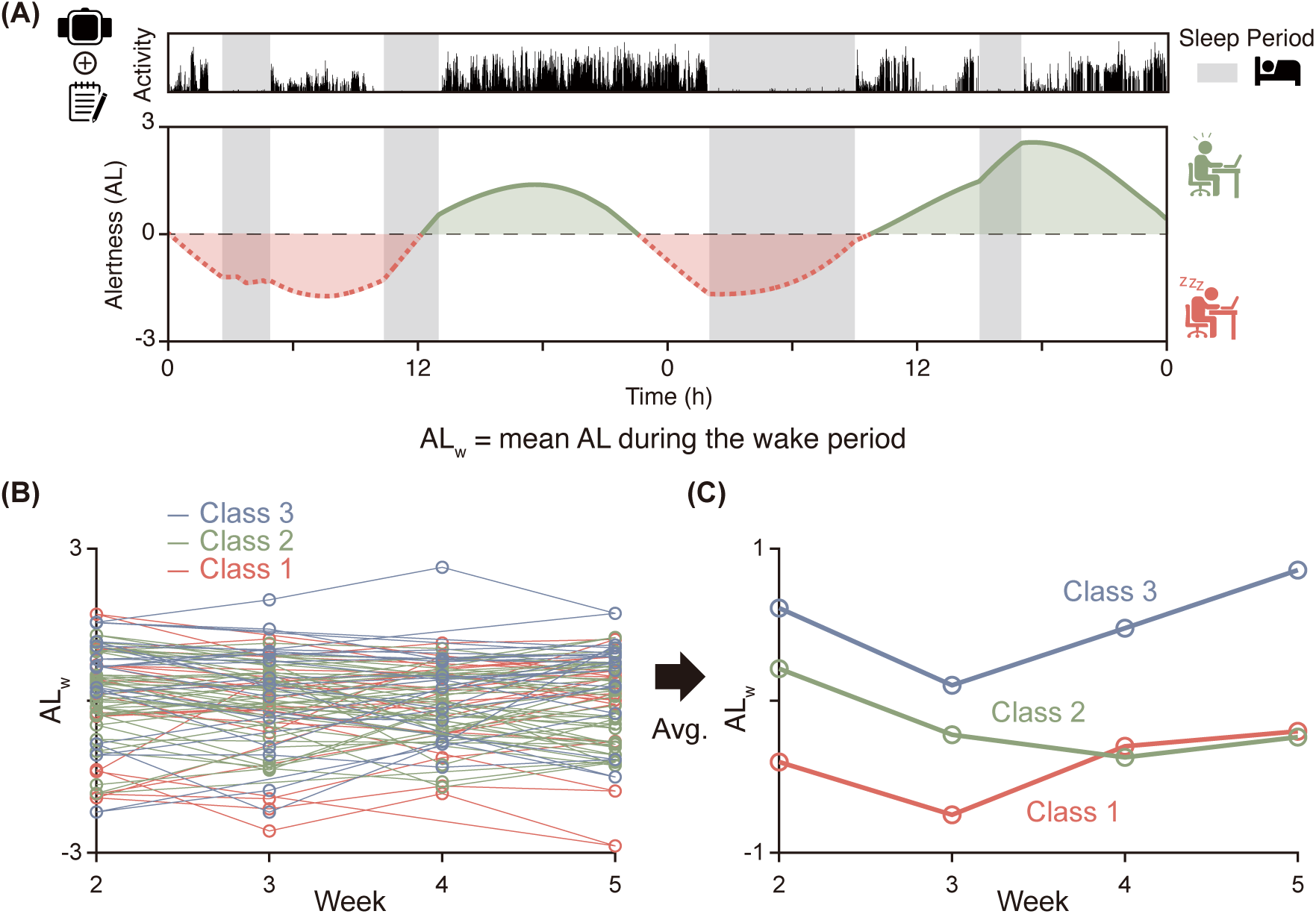
Model-predicted alertness trajectories parallel self-reported sleep quality across identified classes. (A) To objectively assess the intervention’s impact, we used a mathematical model that estimates alertness (AL) from individual sleep patterns (grey region) derived from sleep diaries and wearable devices. AL values above zero (green solid line) indicate effortless waking, whereas values below zero (red dashed line) indicate effortful waking. Weekly mean AL (AL_w_) during wake periods (outside the grey region). (B) Participant-level AL_w_ time series, excluding the first week to mitigate initialization artifacts, were analyzed. (C) The mean AL_w_ trajectory of each class closely resembled self-reported sleep quality, suggesting that the intervention produced measurable improvements in both subjective (sleep quality) and objective (alertness) outcomes.

The individual AL_w_ time series exhibit distinct patterns, akin to SQ (Fig 5B). Upon averaging the time series for each class, the mean AL_w_ trajectories show a similar pattern to that of SQ: steady low level (Class 1), steady moderate level (Class 2), and improvement (Class 3). This suggests that the intervention not only can enhance subjectively perceived sleep quality but also improve physically sensed alertness within participants exhibiting specific conditions identified in the previous section (i.e., Class 3).

## 4. Discussion

### 4.1. Methodological Implications

The GMM, a pivotal analytical framework in this study, has been widely used to characterize the heterogeneity of longitudinal trajectories in clinical investigations. For instance, the GMM was implemented in delineating trajectory classes for psychological constructs such as depression, anxiety, and stress among college students (McLaughlin & King, 2015), as well as cognitive performance trajectories in individuals with hypertension (Zhang & Jiang, 2022). Furthermore, its utility extends to analyzing developmental trajectories of self-reported sleep-related characteristics, such as insomnia or sleep disturbance (Arnison et al., 2022; Van Onselen et al., 2012), similar to the current study. Additionally, GMM serves as a robust tool for assessing the effectiveness of interventions. For example, it has been applied to evaluate exposure-based psychotherapies for individuals with posttraumatic stress disorder (Allan et al., 2017) and internet-based CBTI among those with subclinical depressive symptoms (Batterham et al., 2017). These previous studies support the rationale for utilizing the GMM approach to analyze the effect of intervention on SQ of shift workers, as undertaken in this study.

To complement subjective evaluations of sleep quality, we incorporated a mathematical model for predicting alertness based on objective sleep data. Mathematical models of alertness prediction have been progressively refined and validated since the 1980s (Klerman & St Hilaire, 2007; Mallis et al., 2004). In this study, we applied a recently developed physiologically based model specifically designed to capture dynamic circadian processes under irregular sleep and light-exposure patterns (Hong et al., 2021; Phillips et al., 2010; Skeldon et al., 2017; Song et al., 2023; Swaminathan et al., 2017). Notably, the circadian component of this model has been shown to accurately forecast circadian phase, representing the most effective approach for field applications (Stone et al., 2020). It has also been employed to elucidate causal dynamics between sleep, circadian phase, and mood fluctuations (Song et al., 2024), as well as to predict mood episodes (Lim et al., 2024). Collectively, these prior validations and applications support the use of this model in shift-working populations and provide a robust framework for assessing the alignment of sleep with circadian rhythms under irregular schedules. Future studies may further leverage this model to evaluate shift workers’ sleep and to compare its outputs with other measures of sleep quality, both subjective and objective.

### 4.2. Factors that influence latent trajectories of treatment response

Significant factors distinguishing shift workers whose SQ improved from those did not improve were identified from the latent trajectories of SQ (see section 3.2 for details). The identified factors are consistent with several prior investigations. Specifically, Oh et al. examined 568 individuals, revealing that engaging in fire suppression and emergency duties was associated with a higher likelihood of experiencing poor SQ compared to administrative duties (Oh et al., 2022). Additionally, this study, along with another investigation involving 109 US firefighters, demonstrated that the 3-day cycle was associated with better SQ than other shift work schedules, such as 9- or 21-day cycles (Billings & Focht, 2016; Oh et al., 2022). This suggests that a shift work schedule that maintains relatively consistent sleep patterns is more conducive to improving SQ than a schedule that fluctuates more frequently. This aligns with the findings of the systematic literature review, which indicate that sleep regularity is an important factor in overall health including sleep quality (Sletten et al., 2023). Furthermore, previous studies have indicated significant correlations between longer sleep onset latency or heavy alcohol consumption and poor SQ (Augner, 2011; Devenney et al., 2019). Additionally, consistent with previous studies have shown that insomnia severity was a predictor of treatment response in individuals receiving CBTI (Blom et al., 2021). Previous studies have also shown that alcohol consumption was a risk factor for poor sleep quality, which is consistent with our findings (Ma et al., 2018; Songkham et al., 2018).

### 4.3. Clinical Implications

This study addressed the challenge of assessing sleep quality in shift workers by collecting daily sleep quality and analyzing its trajectory. This approach allows for a more comprehensive understanding of who benefits from the sleep intervention and how to tailor interventions for future use. Similar to non-shift workers, individual differences, especially more severe insomnia symptoms characterized by longer sleep onset latency, were associated with worse outcomes, while less alcohol consumption was associated with improved outcomes. Tailoring intervention durations according to symptom severity, rather than using a uniform approach, could optimize resource allocation for enhanced care in improving sleep among shift workers. Additionally, organizations with high stress, such as firefighters, who must always be prepared to respond to the next emergency, are more vulnerable to using alcohol or other substances with the purpose of sleep induction (Zegel et al., 2019). Sleep education on the effects of alcohol on sleep quality, in addition to screening for alcohol use prior to treatment and monitoring use over time, may also improve treatment response.

Work-related factors may need to be addressed on both individual and organizational levels. On an individual level, differentiating sleep interventions based on the role of the patient within the organization may be important. For example, a firefighter who is working on the emergency response team may be woken up several times a night when they are on call, with a higher likelihood of sleep disturbance compared to a firefighter who works in fire suppression, who may be able to sleep through the night more frequently if no fire has occurred. On an organizational level, since CBTI is often conducted on an individual referral basis, organizational factors that contribute to sleep difficulties may interfere with treatment and be a major contributor to sleep disturbance but are not adequately addressed. Clinicians working with shift workers may need to also act as advocates on behalf of their patients to emphasize the importance of sleep on performance and well-being, as well as communicate optimal shift cycles as organizations may not be aware or less educated about circadian factors that interrupt sleep.

### 4.5. Limitations

Nevertheless, there are certain limitations to this study. First, the study variables relied on participant self-report, which may indicate a self-report bias in the observed relationships. To address this limitation, we integrated a multi-method approach to validate these findings using model-predicted alertness based on objectively observed sleep-wake data.

Second, the sample size of this study was small due to the nature of the shift worker population and the multi-period follow-up intervention program. The participants in this study took part in a long-term intervention program that included daily sleep diaries and actigraphy. It is challenging and expensive to obtain large samples for such field studies, and most are conducted on a small scale for practical reasons of restraints with cooperation, time and economic reasons. Whereas many latent variable studies, including GMMs, are typically conducted on multidimensional variables with relatively large sample sizes due to the number of parameters and their identifiability and estimability. However, a sample size as small as 30 per subgroup is known to be sufficient when the subgroups of the latent class are well separated, and normality is satisfied (Lubke & Luningham, 2017; Nylund-Gibson & Choi, 2018). Shi et al. also showed that latent growth models can produce meaningful results even with small sample sizes (n<100) (Shi et al., 2021). It is important to note that the sample size used in GMM is not simply the number of participants, but rather all observed time points provided by each participant over multiple periods. Therefore, while the number of participants may be small, the comprehensive longitudinal information used in the analysis ensures that the model’s identifiability and estimability are satisfied. Despite the small sample size, this study contributes to identifying distinct latent subgroups based on SQ trajectories during the intervention period, which can inform interventions to improve the sleep, well-being, and performance of shift workers. GMM allowed us to uncover common trajectories, and through multinomial logistic regression, we examined how these sleep patterns relate to various factors. The combined analysis offers significant implications for interventions, demonstrating that even with limited data, valuable insights can be derived.

Third, this study only included shift-working firefighters, which limits the generalizability of our findings to this specific occupation. Additionally, since no specific exclusion criteria except non-shift work for the study participants were defined, the results may have been influenced by preexisting sleep disorders and psychiatric disorders. Furthermore, most participants in this study were male (88%), which may have restricted the influence of gender on the SQ trajectories. Therefore, careful consideration should be given to the generalization of the results of this study. Furthermore, including more diverse samples of shift workers would be meaningful for future research.

Fourth, the mathematical model used to predict alertness relies on uniform, population-level parameters, which fail to account for the reported inter-individual differences (Van Dongen et al., 2005). Prior research has highlighted the importance of individualizing models that predict alertness (Knock et al., 2021; Song et al., 2023). Future studies could explore this individualization, potentially enhancing the effectiveness of objective assessments of sleep quality.

Finally, the number and variety of predictors and outcomes were limited in this study, mainly due to the sample size. Future research with sufficient sample size could broaden the scope to encompass a more comprehensive array of predictors and outcomes, thereby reducing the risk of omitted variable bias stemming from potential confounding factors, such as insomnia characteristics and resilience.

## 5. Conclusion

This study emphasizes that latent patterns of treatment response are influenced not only by an individual’s sleep characteristics but also by work-related variables, such as shift duration and type of shift work. This underscores the importance of personalized and targeted interventions that address the specific needs and challenges of shift workers. By considering these factors, clinicians can develop more effective strategies. While individual-level factors can be addressed with interventions like CBTI, organizational-level changes may also be necessary to improve the sleep of shift workers. By collectively addressing these factors, it becomes possible to develop more effective, tailored interventions to enhance sleep and well-being in shift workers.

## 6. Data Availability Statement

The data that support the findings of this study are available from the corresponding author upon reasonable request.

## 8. Declaration of generative AI and AI-assisted technologies in the writing process

During the preparation of this work the author(s) used GPT4.0 in order to assist with spelling and grammar checks. After using this tool/service, the authors reviewed and edited the content as needed and take full responsibility for the content of the publication.

## Appendix A: Intervention Details

The FIT-INdividual intervention was developed based on cognitive-behavioral therapy for insomnia, and expanded upon a group-based sleep intervention for Korean firefighters that was based on brief-behavioral therapy for insomnia (Jang et al., 2020). FIT-Individual was modified to incorporate elements specifically for shift workers and was adapted into a telehealth format. The intervention consisted of four 50-minute weekly sessions (Table A.1), all led by licensed clinical psychologists. In addition, participants completed daily sleep diaries after waking up, which were collected online, and the research assistant monitored whether they were submitted daily.

**Table A.1.** Components for each session of the Firefighter’s Therapy for Insomnia and Nightmares-INdividual version (FIT-INdividual)

| Session | Treatment Module | Goals and Activities | Homework |
| --- | --- | --- | --- |
| 1<sup>st</sup> session | Sleep Education and Sleep Hygiene | • Explaining definitions of insomnia and shift work disorder<br>• Introduced guidelines on sleep hygiene<br>• Select a sleep schedule<br>• Guide the sleep diary | Sleep Diary |
| 2<sup>nd</sup> session | Stimulus Control and Relaxation Circadian rhythm intervention | • Guide a sleep-friendly environment<br>• Provide a sleep guideline<br>• Demonstrate relaxation skills | Sleep Diary |
| 3<sup>rd</sup> session | Exploring Dysfunctional Thoughts that maintain Insomnia | • Exploring the dysfunctional beliefs<br>• Conduct cognitive restructuring | Sleep Diary |
| 4<sup>th</sup> session | Relapse Prevention | • Evaluated the entire previous intervention<br>• Discuss relapse prevention strategies<br>• Summary |  |

The first session conducted sleep education, which involved explaining insomnia, shift work disorder, the principles of sleep (the two process model), and the theoretical basis of the development of insomnia (the 3-P model). Guidelines for sleep hygiene tailored for shift workers were also introduced, and a sleep schedule was prescribed.

The second session included stimulus control techniques, during which participants were instructed to create a sleep-friendly environment. A circadian rhythm component was added to the intervention to address participants’ needs (e.g., avoiding sunlight by wearing light-blocking sunglasses while returning home from the night shift and using dark curtains while sleeping after the night shift).

The third session consisted of cognitive therapy techniques. During this session, participants identified dysfunctional beliefs about their sleep and were guided through cognitive restructuring techniques to modify thoughts interfering with their sleep.

The final session was focused on evaluating the intervention and discussed relapse prevention. Participant and clinician discussed achieves made and changes in sleep patterns during the intervention. Additionally, the clinician explained how insomnia can emerge in situations of high stress, and introduced stress inoculation techniques by discussing high-risk situations and responses. Participants submitted sleep diaries until one week after the last session to monitor their sleep after the intervention.

## Appendix B: Measures and Preprocessing Details

### B.1. Measures

#### Demographic and lifestyle information

Participants completed a self-report questionnaire about demographic and lifestyle information prior to intervention. The questionnaire includes individual factors such as gender, age, height, weight and daily average amount of caffeine intake. It also includes work-related factors such as primary duty, rank, length of service, shift work period and shift schedule. Shift schedule cycles can be either 3-day cycles or 21-day cycles: A 3-day cycle consists of 24 hours on, followed by 48 hours off, while a 21-day cycle consists of consecutive day shifts, alternating night shifts, 24 hours on, followed by 24 hours off.

#### Insomnia Severity Index (ISI)

The ISI is a self-report questionnaire comprising seven items that assess insomnia symptoms experienced during the preceding month (Bastien et al., 2001). Each item is rated on a 5-point Likert scale (0-4), with a total score ranging from 0 to 28. A higher score indicates more severe insomnia symptoms. The Korean version of the ISI was validated by Cho et al. (Cho et al., 2014). Internal consistency (Cronbach’s alpha) in this study was 0.75.

#### Fatigue Severity Scale (FSS)

The FSS is a self-report questionnaire comprising nine questions about fatigue in the week prior to reporting (Krupp et al., 1989). Each item is rated on a 7-point Likert scale, and the total score is calculated as the average of the scores for each item, with a range of 1 to 7 points. The Korean version of the FSS was validated by Lee et al. (Lee et al., 2013). The internal consistency (Cronbach’s alpha) in this study was 0.88.

#### Shift Work Disorder Questionnaire (SWDQ)

The SWDQ is a questionnaire that assesses insomnia symptoms and daytime sleepiness related to shift work in the past month (Barger et al., 2012). It consists of four questions about sleep problems during shift work, well-being while awake, likelihood of falling asleep at work, and likelihood of falling asleep while driving. The questionnaire is used to identify at-risk individuals for shift work disorder. Internal consistency (Cronbach’s alpha) in this study was 0.61.

#### Sleep diary

Participants in the intervention were instructed to complete weekly sleep diaries, recording their sleep from the previous night upon waking. The sleep diary was based on the Consensus Sleep Diary (CSD) (Carney et al., 2012). It included questions about various aspects of sleep, such as total sleep time (TST), number of awakenings (NWAK), sleep onset latency (SOL), sleep efficiency (SE), wake after sleep onset (WASO), nap, and sleep quality (SQ). SQ was evaluated subjectively through self-report. Each day, participants rated their SQ for the previous night’s sleep, which was reported on a scale of 1 (poor) to 5 (excellent). Additionally, alcohol intake and whether dispatches occurred during sleep periods on night shifts were recorded. For sleep segments interrupted by dispatches, data were collected separately, with WASO excluding dispatch time. Diary data was collected over a period of approximately five weeks, spanning from the week prior to the initial session to the week succeeding the final session.

#### Actigraphy

All participants wore actigraphy devices (Readiband V5, Fatigue Science Inc., Vancouver BC, Canada) to measure objective sleep while participating in the study. Actigraphy is a wrist-worn activity monitor capable of sensing movement and measuring sleep data. The device used in our study (Readiband) has been validated against polysomnography, the gold standard in sleep assessment, and compared to other sleep monitoring devices (Chinoy et al., 2021). Sleep indices about sleep duration and sleep timing, such as sleep onset and wake time, calculated from the automatic algorithm within the device (Russell et al., 2000), were collected.

### B.2. Data pre-processing

Exploratory analyses were conducted using demographic and lifestyle information and data collected from questionnaires and sleep diaries to identify outliers and conduct a preliminary investigation about distributional disparities and/or correlations with self-reported sleep quality. Subsequently, essential variables were derived and organized from the demographic and lifestyle data. For instance, Body Mass Index (BMI) was computed from height and weight, and certain categorical levels were amalgamated to ensure suitable sample sizes for analysis. Specifically, primary duties, initially categorized into emergency, fire, rescue, and other work-related factors, were consolidated for analytical purposes, grouping all non-emergency categories under "other." Similarly, ranks from Firefighter to Fire Chief were reclassified into a binary level (firefighter vs. other). Additionally, average daily caffeine intake was recategorized into three levels (none vs once a day vs twice or more per day). These recategorizations were implemented because the sleep quality associated with each original category showed no statistically significant differences.

Internal consistency of the pre-intervention clinical indicators, ISI, FSS, and SWDQ, were evaluated. Additionally, the unidimensionality of their first principal components, explaining 91.4%, 83.4%, and 95% of the variance, respectively, further supported their reliability. Hence, averaging individual item responses was deemed suitable for capturing firefighters’ baseline levels of insomnia, fatigue, and shift work related stress. Sleep diaries recorded sleep parameters for up to four distinct sleep episodes each day, segmented by dispatch. From these parameters, the average sleep onset latency was computed, while the total sleep time, time in bed, number of wake episodes, and wakefulness after sleep onset were summed. Additionally, sleep efficiency was calculated by dividing the total sleep time by the time in bed.

Minute-by-minute sleep data obtained using actigraphy offered intricate insight into wakefulness periods during each sleep episode, a level of detail not captured by sleep diaries. This data is utilized for simulating a mathematical model to predict alertness, relying on continuous and detailed sleep-wake patterns (see **Appendix C.3** for detailed methodology). In cases where wearable data were unavailable but sleep diary information was accessible, missing values were imputed based on sleep diary data, following previous studies utilized the mathematical model (Hong et al., 2021; Song et al., 2023). Specifically, missing wearable data such as sleep onset and offset were imputed from participants’ reported sleep diaries and then interpolated to the minute. If sleep diary data was missing within a week from the start or end of the intervention, that period was excluded from model simulations. If missing data persisted for over two consecutive days during other intervals within the intervention period, participants were excluded from the analysis. This criterion led to the exclusion of two participants. For a single day, where the missing data persisted for only one day during other intervals and the surrounding five bedtimes were relatively regular (SD of sleep bedtime onset and offsets were 28.8 minutes and 7.2 minutes), the sleep period was estimated based on the previous days’ records. Consequently, about 7% of the total minute epochs were imputed.

## Appendix C: Analysis Details

### C.1. Time series clustering using growth mixture model

Our main interest was to investigate whether and to what extent participants’ sleep quality is affected by the intervention, even in the context of their demanding shift work schedules. This was implemented by examining individual levels of sleep quality for common trends over the intervention period. However, each individual attended each intervention session, according to their own available schedule, resulting in variations in both the timing and duration of the intervention sessions. Consequently, this variability posed a challenge in identifying underlying latent trajectories regarding the effectiveness of the interventions. To overcome this challenge, a representative value of sleep quality of each intervention session was calculated using daily reported SQ from participants’ sleep diaries, allowing common trajectories to be identified.

GMM was then used to identify latent trajectory classes of weekly average SQ across five time points. Unconditional models with one to five classes for SQ were tested, and parameters for intercepts, linear, and quadratic slopes were estimated for each latent class. Full information maximum likelihood (FIML) estimation was used to handle missing data (Giletta et al., 2015). The optimal number of latent classes was selected according to the following criteria (Nylund et al., 2007): lower information criteria fit indices (AIC, BIC, SSBIC), higher entropy values (>0.80), likelihood ratio tests (LRTs; p < 0.05 indicates better fit for c classes than c-1), and ensuring at least 5% of participants in each class.

### C.2. Latent class trajectory prediction using multinomial logistic regression

Once the number of latent classes has been identified, it becomes important to understand the sources of these differences in trajectories. For the number of latent classes greater than 2, multinomial logistic regression can be used to examine the risk factors associated with trajectory membership and to predict the likelihood of latent class membership given the risk factors. Multinomial logistic regression was performed for the 5-week latent classes (n=243) using collected predictors, including six categorical and eight continuous demographic, lifestyle, and clinical variables, in addition to six sleep parameters. Categorical variables include gender (Gender; M vs F), age (Age; <30, 30-39, 40+), primary duty (Duty; emergency vs other), fire department rank (Rank; fire fighter vs others), shift schedule (Shift schedule; 3-day cycle vs 21-day cycle), average daily caffeine intake (Caffeine; none vs once a day vs twice or more per day). Continuous variables include total shift work period (Shift work period; years), total length of service (Len. of Service; years), number of dispatch days (Dispatch) and alcohol intake days (Alcohol) per week, and averaged item responses of ISI (ISI; 0∼4), FSS (FSS; 0∼6), and SWDQ (SWDQ; 0∼3) during the pre-experiment period. Sleep parameters include weekly averaged total sleep time (TST; min), sleep efficiency (SE; %), number of awakenings (NWAK; times), wakefulness after sleep onset (WASO; min), sleep onset latency (SOL; min), and number of naps (Nap; times).

To avoid multicollinearity, backward elimination was used as variable selection for logistic analyses. Data pre-processing, preliminary exploratory data analysis and analyses for GMM and latent class trajectory prediction using multinomial logistic regression were performed using SAS 9.4 and R 4.3.2.

### C.3. Alertness prediction using a mathematical model

For an objective assessment of the intervention effect, we predicted individual alertness (AL) using a validated mathematical model (Hong et al., 2021; Phillips et al., 2010; Skeldon et al., 2017; Song et al., 2025; Song et al., 2023; Swaminathan et al., 2017) (see Supplementary information of (Hong et al., 2021) for model details). The model simulates temporal changes in AL based on individual sleep patterns, with AL values below zero (red dashed line in Fig 4A) indicating effortful wakefulness and values above zero (green solid line in Fig 4A) indicating effortless wakefulness. This approach has previously been shown to predict daytime sleepiness and momentary alertness in shift-working nurses (Hong et al., 2021; Song et al., 2023). Furthermore, a sleep-scheduling intervention derived from this model has been demonstrated to improve alertness in shift workers (Song et al., 2025).

We simulated this model based on individual sleep and light exposure patterns measured using actigraphy and sleep diaries (see **Appendix B.3** for details). Specifically, the light exposure pattern was deduced from the sleep diary data, assuming 0 lux from the reported light off time for sleep to wake time, 250 lux during 8:00∼20:00 hours, and 40 lux otherwise during wake periods (Knock et al., 2021; Song et al., 2023). The initial condition was determined by assuming that individuals followed the same pattern observed during the initial week, repeated five times. The model simulations were performed with ode15s solver in MATLAB 2021a software (Natick, MA, USA).

